# Head-to-head comparison of composite and individual biomarkers to predict clinical benefit to PD-1 blockade in Non-Small Cell Lung Cancer

**DOI:** 10.1101/2023.10.19.23297261

**Authors:** Karlijn Hummelink, Vincent van der Noort, Mirte Muller, Robert D. Schouten, Michel M. van den Heuvel, Daniela S. Thommen, Egbert F. Smit, Gerrit A. Meijer, Kim Monkhorst

**Affiliations:** Department of Pathology, Division of Diagnostic Oncology; Department of Thoracic Oncology, Division of Medical Oncology; Department of Biometrics, Netherlands Cancer Institute, Amsterdam, The Netherlands; Department of Pulmonary Diseases, Radboud University Medical Center, Nijmegen, The Netherlands; Division of Molecular Oncology and Immunology, Netherlands Cancer Institute, Amsterdam, The Netherlands; Department of Pulmonary Diseases, Leiden University Medical enter, Leiden, The Netherlands

## Abstract

**Background:** Treatment with PD-(L)1 blocking agents has demonstrated durable efficacy in advanced NSCLC, but only in a minority of patients. Multiple biomarkers for predicting treatment benefit have been investigated, but their combined performance has not been extensively studied. Here, we assess the combined predictive performance of multiple biomarkers in a series of NSCLC patients treated with nivolumab.

**Methods:** Pretreatment samples from 135 patients treated with nivolumab were used to assess the predictive performance of CD8 tumor-infiltrating lymphocytes (TILs), intratumoral (IT) localization of CD8 TILs, PD-1 high expressing TILs (PD1^T^ TILs), CD3 TILs, CD20 B-cells, tertiary lymphoid structures (TLS), PD-L1 tumor proportion score (TPS) and the Tumor Inflammation score (TIS). Patients were assigned to a training (n=55) and validation set (n=80). The primary outcome measure was Disease Control at 6 months (DC 6m) and the secondary outcome measure was DC at 12 months (DC 12m).

**Results:** In the validation cohort, the two best performing composite biomarkers (i.e. CD8+IT-CD8 and CD3+IT-CD8) demonstrated similar or lower sensitivity (64% and 83%) and NPV (76% and 85%) than the individual biomarkers PD-1^T^ TILs and TIS (sensitivity: 72% and 83%, NPV: 86% and 84%) for DC 6m, respectively. Also, at 12 months, both selected composite biomarkers (CD8+IT-CD8 and CD8+TIS) showed less predictive performance compared to PD-1^T^ TILs and TIS alone. PD-1^T^ TILs and TIS showed high sensitivity (86% and 100%) and NPV (95% and 100%) for DC 12m. PD-1^T^ TILs could better discriminate patients with no long-term benefit, since specificity was substantially higher as compared to TIS (74% versus 39%).

**Conclusion:** Composite biomarkers did not show improved predictive performance compared to PD-1^T^ TILs and TIS alone for both the 6- and 12-months endpoint. PD-1^T^ TILs and TIS identified patients with DC 12m with high sensitivity. Patients with no long-term benefit to PD-1 blockade were most accurately identified by PD-1^T^ TILs.

## Introduction

The success of monoclonal antibodies targeting the inhibitory receptor programmed cell death protein 1 (PD-1) and its ligand programmed death-ligand 1 (PD-L1) has changed the treatment landscape of advanced stage non-small cell lung cancer (NSCLC). A subset of patients treated with these PD-(L)1 blocking agents can achieve durable responses and gain a large survival benefit(1–7). Unfortunately, the majority does not derive durable clinical benefit, highlighting the need for predictive biomarkers to support treatment decision making in clinical practice. Specifically, biomarkers to exclude patients who are unlikely to benefit from PD-1 blockade therapy can offer patients alternative treatment options.

Tumor PD-L1 expression, as detected by immunohistochemistry (IHC), has been studied as a predictive biomarker in multiple clinical trials(8). A positive correlation between PD-L1 expression and treatment outcome has been reported in advanced stage NSCLC patients(1,5–7). However, approximately 60% to 70% of patients with PD-L1 positive tumors do not respond(1,2,5). Besides this, PD-L1 assessment by IHC is hampered by intratumor heterogeneity, interassay- and interobserver variability as well as pre-analytical variation(9–14). Tumor Mutation Burden (TMB), reflecting the number of somatic mutations as a surrogate of potential tumor antigenicity, has also shown predictive potential but clinical implementation remains challenging due to the lack of a robust and predictive TMB cut-off and the technical issues that arise due to variation across platforms(15–17).

For these reasons there is an urgent need for biomarkers that can more accurately predict response to PD-(L)1 blockade in advanced NSCLC. Since PD-(L)1 blockade is thought to reinvigorate tumor-reactive T cells(18–20), several T cell markers have been investigated. For example, the density of CD8^+^ tumor infiltrating lymphocytes (TILs) has been correlated to response to PD-(L)1 blockade in melanoma(18), colorectal cancer(21), and NSCLC(22,23). In previous work we have shown that a distinct T cell population, termed PD-1^T^ TILs, can predict clinical benefit in NSCLC(24,25). Notably, these PD-1^T^ TILs predominantly localize in tertiary lymphoid structures (TLS)(24). B cells, which are a critical component of these TLS, have also been linked to response to PD-(L)1 blocking agents(26–28). Other studies developed predictive RNA expression signatures, such as the “tumor inflammation signature” (TIS), to characterize features of immune activity in the tumor microenvironment (TME)(29–31).

Although all of these biomarkers have shown a certain predictive potential, their accuracy is still limited which is presumably caused by multiple components that are involved in the antitumor immune response. Hence, combining biomarkers could potentially improve their predictive accuracy, as previously has been shown for the combination of TMB with PD-L1(32,33) and CD8 TILs with PD-L1(22,34). Therefore, the aim of the present study was to investigate the performance of CD8, PD-1^T^ TILs and CD3 TILs, CD20^+^ B cells, TLS, PD-L1 and TIS as pairs of biomarkers, compared to single biomarkers, for prediction of clinical benefit to PD-1 blockade in NSCLC.

## Methods

### Patients, endpoints and samples

In this study, 162 patients with pathologically confirmed stage IV NSCLC were eligible for efficacy analysis. All patients started second or later line monotherapy nivolumab, 3mg/kg as an IV infusion every two weeks for at least one dose, between October 2014 and August 2017 at the Netherlands Cancer Institute/Antoni van Leeuwenhoek hospital (NKI-AVL), The Netherlands. Patients with tumors harboring known sensitizing EGFR mutations or ALK translocations were excluded from treatment. Patients were randomized into a training and validation cohort. Randomization was stratified by treatment outcome at 6 months and at 12 months. Since we could only generate gene expression data in 68/162 (42%) of patients’ tumors, additional stratification was done by whether mRNA expression analysis was performed or not. Stratification for missing values of other biomarkers was not performed, as the number of excluded samples per biomarker was low (range 1 to 32) (see **Supplementary Fig. S1** and later in this section).

Response was assessed per Response Evaluation Criteria in Solid Tumors (RECIST) version 1.1. Patients with progressive disease (PD) who were not evaluable for response were determined by the treating physician as PD. The primary clinical outcome was Disease Control (DC) (complete response (CR)/partial response (PR) or stable disease (SD)) at 6 months following initiation of treatment. DC 12m (CR/PR/SD that lasted ≥12 months) was used as secondary outcome measure to predict long-term efficacy to PD-1 blockade.

Pretreatment formalin-fixed paraffin embedded (FFPE) tumor tissue samples were collected from all patients. Written informed consent was obtained from all patients for research usage of material not required for diagnostic use by institutionally implemented opt-out procedure. The study was conducted in accordance with the Declaration of Helsinki. The data was accessed for research purposes after the approval of the Institutional Review Board (IRB) of the Netherlands Cancer Institute on January 11, 2018 (CFMPB586). After K.H., M.M., R.D.S., M.M.H., E.F.S. and K.M. retrieved archived tumor samples and response data from medical records, all patients were pseudonymized. PD-1^T^ TIL and PD-L1 tumor proportion score (TPS) data for 94 samples were used from our previous work as well as tertiary lymphoid structures (TLS) and CD20^+^ B cell data for 91 samples(25). In 27 patients, none of the biomarkers could be assessed because samples did not contain tumor tissue. In one sample no tumor tissue was left for CD8 and PD-1^T^ TIL analysis, as well as in five samples for CD3 TIL, TLS and CD20^+^ B cell analysis **(Supplementary Fig. S1)**. An additional number of 32 patients were excluded for PD-1^T^ TIL analysis based on the following criteria: samples contained less than 10,000 cells (n=12), were obtained from endobronchial lesions (n=16), contained abundant normal lymphoid tissue (n=1) and showed fixation and/or staining artefacts (n=2) **(Supplementary Fig. S1)**. As described before, we excluded bronchial biopsies because they frequently showed unspecific antibody staining due to mechanical damage, and lymph node resections due to presence of PD-1^+^ T cells in normal abundant lymphoid tissue, which could potentially lead to false positive results(25). One sample was excluded for CD8 TIL, CD3 TIL, TLS, CD20^+^ B cell and PD-L1 analysis because of fixation/staining artefacts. One sample contained less than 2,000 cells and was excluded for CD8 TIL, CD3 TIL, TLS and CD20^+^ B cell analysis. 67 patients (41%) were excluded for mRNA expression analysis because of low RNA yield and/or low RNA quality **(Supplementary Fig. S1)**.

### Immunohistochemistry

CD8 immunostaining of samples was performed on a BenchMark Ultra autostainer Instrument (Ventana Medical Systems) on 3 µm paraffin sections from FFPE blocks. Sections were initially baked at 75°C for 28 minutes and deparaffinised in the instrument with EZ prep solution (Ventana Medical Systems). Heat-induced antigen retrieval was carried out using Cell Conditioning 1 (CC1, Ventana Medical Systems) for 32 minutes. CD8 was detected using clone C8/144B (1/200 dilution, 32 minutes at 37°C, Agilent/DAKO). Bound antibody was detected using the OptiView DAB Detection Kit (Ventana Medical Systems). Slides were counterstained with Hematoxylin and Bluing Reagent (Ventana Medical Systems).

PD-1 immunostaining was detected using clone NAT105 (Roche Diagnostics), PD-L1 using clone 22C3 (Agilent/DAKO) and CD68 using clone KP1 (Agilent/DAKO). For the double staining CD20 (yellow) followed by CD3 (purple) we used clone L26 (Agilent/DAKO) (CD20) and clone SP7 (Thermo Fisher) (CD3). All immunostainings were performed as described previously(25).

CD8, PD-1, PD-L1 and CD68 immunostainings were scanned at x20 magnification with a resolution of 0.50 per µm^2^ using an Aperio slide AT2 scanner (Leica Biosystems). CD20-CD3 immunostainings were scanned at x20 magnification with a resolution of 0.24 per µm^2^ using a 3Dhistech P1000 scanner. PD-L1 and CD68 data were uploaded on Slide Score, a web platform for manual scoring of digital slides using a scoring sheet (www.slidescore.com).

### Digital quantification of CD8 and PD-1^T^ TILs

Digital image analysis was performed by a trained MD (K.H.) and supervised by an experienced pathologist (K.M.) using the Multiplex IHC v1.2 module from the HALO^TM^ image analysis software, v2.3.2089.69 (Indica Labs). Researchers were blinded for clinical outcome. Classification of CD8 lymphocytes on single stains was performed using a computationally derived cut-off of 0.3 optical density (OD), which reflects the intensity of the staining. This cut-off was identified by manually optimizing the detection of CD8 positive stained cells in FFPE samples. An image analysis algorithm utilizing a cut-off of 0.3 OD was generated for automated analyses of CD8 lymphocytes in subsequent FFPE samples. The quantification of PD-1^T^ TILs was performed as described previously(25).

The number of CD8 and PD-1^T^ TILs per mm^2^ tumor area were determined. Tumor areas were digitally annotated as described previously(25). PD-1^T^ TIL data of 94 samples were used from previous work(25). For regional analysis of CD8 lymphocytes, classifiers were trained to identify stromal and tumoral regions in which the CD8 lymphocytes were quantified separately. The percentage CD8 lymphocytes in tumoral regions (i.e. intra-tumoral (IT)) compared to total CD8 TILs was calculated **(Supplementary Table S1)**.

### Scoring of tertiary lymphoid structures

The HALO^TM^ image analysis software, v2.3.2089.69 (Indica Labs) was used to determine the number of TLS and the combined number of TLS and lymphoid aggregates (TLS+LA) per mm^2^ tumor area on a CD20-CD3 double immunostaining as described previously(25). TLS and TLS+LA data of 91 samples were used from previous work(25) **(Supplementary Table S1)**.

### CD20 and CD3 quantification by digital image analysis

The total area with CD20 expression was measured using a previously generated image analysis algorithm from the Area Quantification v1.0 module of HALO^TM^ image analysis software (Indica Labs)(25). The same algorithm was used to measure the total area with CD3 expression. The CD20-positive and CD3-positive area were normalized per mm^2^ tumor area. Cell numbers were not quantified as no reliable algorithm could be established due to dense clustering of CD20^+^ or CD3^+^ cells in and at the border of TLS. Tumor areas were digitally annotated as described previously(25). CD20 data of 91 samples were used from previous work(25) **(Supplementary Table S1)**.

### PD-L1 scoring

PD-L1 TPS was determined using the qualitative, clinical grade LDT IHC assay (22C3 Agilent/DAKO) as described previously(25). PD-L1 TPS data of 94 samples were used from previous work(25) **(Supplementary Table S1)**. The CD68 staining was compared to the PD-L1 staining to exclude macrophages that are both CD68^+^ and PD-L1^+^ which can introduce false positive results.

### RNA extraction and hybridization to nCounter tagset

RNA of pretreatment FFPE samples from the NKI-AVL cohorts were isolated with the AllPrep DNA/RNA FFPE isolation kit (#80234, Qiagen) according to the instructions of the manufacturer and quantified by Tapestation (Agilent). 200 to 300 ng RNA were hybridized to Nanostring PanCancer IO 360 Panel code set (Nanostring), according to the recommendations of the manufacturer. After hybridization non-bound probes were washed off and the RNA-probe complex was bound to the cartridge on the Nanostring Flex Prep Station according to manufacturing protocol. The cartridge was sealed and transferred to the Digital Analyzer for imaging.

### Statistical analysis

Patient characteristics were descriptively reported using mean ± SD, interquartile range (IQR) or frequencies (percentages). The Mann-Whitney test for continuous data, Fisher’s exact test for categorical data and linear-by-linear association test for ordinal data were used to assess differences in patient characteristics between cohorts (training and validation) and between outcome groups (disease control vs PD). Differences were considered statistically significant if \**P*<0.05, \*\**P*<0.01, \*\*\**P*<0.001 or \*\*\*\**P*<0.0001.

Genes in the Tumor Inflammation Signature (TIS) are normalized using a ratio of the expression value to the geometric mean of the housekeeper genes used only for the TIS signature and then followed by log2 transformation. The TIS score was calculated as a weighted linear combination of the 18 gene expression values(29,35) **(Supplementary Table S1)**. This analysis was performed by Nanostring as part of their intellectual property.

In the training cohort, univariate models and bivariate logistic models for DC 6m and DC 12m of treatment were constructed using CD8 TILs, IT-CD8 T cells, PD-1^T^ TILs, CD3 TILs, TLS, TLS+LA, CD20^+^ B cells, PD-L1 and TIS. The bivariate models included an interaction term. The bivariate logistic model produces for each patient a number between 0 and 1, reflecting the probability (according to the model) of patients reaching DC 6m or DC 12m. Calculation of the area under the receiver operating characteristic (ROC) curve was used as a measure of discriminatory ability. The predictive performance of different individual and composite biomarkers on the same patient population was described in terms of sensitivity, specificity, positive predictive value (PPV) and negative predictive value (NPV) and compared using the McNemar test. A point on the ROC curve matching a sensitivity of 90% for DC 6m and 90% for DC 12m was selected to calculate corresponding specificity, NPV and PPV. We further aimed for an NPV of ≥90% and a specificity of ≥50%.

Two (closely related) non-parametric approaches were considered to obtain 90% sensitivity for predicting DC 6m and DC 12m from two biomarkers: by choosing a cut-point for each of the two biomarkers and declaring the patient positive (i.e. likely to respond to PD-1 blockade) when either at least one (first method) or both (second method) biomarker values were above their respective cut-point values. The specificities obtained were either equal or worse to those obtained by the parametric method described above (i.e. via logistic regression). Therefore, these non-parametric methods were not used in this study.

Four training models were selected with a cut-off that showed the highest specificity and NPV at the prespecified sensitivities for prediction of DC 6m and DC 12m. This cut-off was used to determine sensitivity, specificity, NPV and PPV in the validation cohort.

## Results

### Biomarker characteristics and demographics

To assess the predictive performance of multiple biomarker combinations we first analyzed 162 pretreatment tumor samples from 162 advanced stage NSCLC patients treated with nivolumab. In total we evaluated 9 biomarkers: (1) the total number of CD8 TILs per mm^2^, (2) the percentage intra-tumoral (IT) CD8 T cells of total CD8 TILs, (3) the number of PD-1^T^ TILs per mm^2^ (4) the CD3-positive area per mm^2^ to estimate the presence of CD3 TILs (5) the CD20-positive area per mm^2^ to estimate the presence of B cells (6) the number of TLS and (7) the combined number of TLS and LA (referred as TLS+LA) per mm^2^, (8) the PD-L1 Tumor Proportion Score (TPS) and (9) the TIS score (NanoString**) (Fig. 1).** We could successfully assess CD8 TILs and IT-CD8 T cells in 132/162 (81%), PD-1^T^ TILs in 103/162 (64%), CD3 TILs, CD20^+^ B cells, TLS and TLS+LA in 128/162 (79%), PD-L1 TPS in 134/162 (83%) and TIS in 68/162 (42%) samples **(Table 1, Supplementary Fig. S1)**. Sample exclusion criteria are shown per biomarker in **Supplementary Fig. S1**.

**Figure 1.**
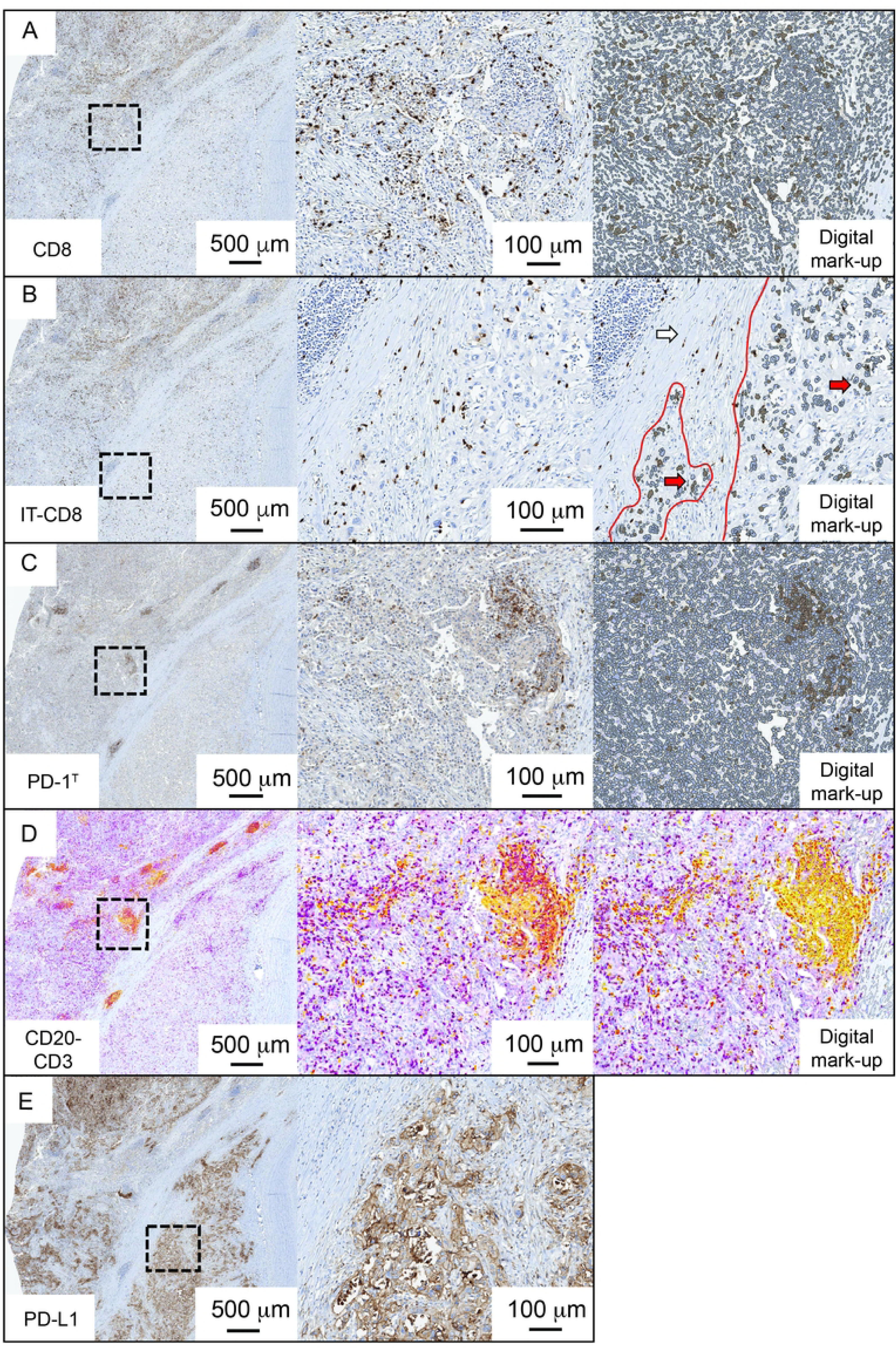
Immunohistochemical analysis of all biomarkers and digital mark-up. **(A)** The left image shows an example of a CD8 immunohistochemical staining (IHC). The black square indicates the area that is shown in the central image. The right image shows the digital markup with CD8 TILs (in brown) and all other cells (in grey). **(B)** The left image shows the same example as shown in A. The black square indicates the area that is shown in the central image. The right image shows regional analysis of only intratumoral (IT) CD8 TILs. Stromal CD8 TILs are not quantified. Red lines indicate the tumor region. Red arrows indicate IT-CD8 TILs. White arrow indicates the area with stromal CD8 TILs. **(C)** The left image shows an example of a consecutive slide stained for PD-1 IHC. The black square indicates the area that is shown in the central image. The right image shows the digital markup with PD-1^T^ TILs (in brown) and all other cells (in grey). **(D)** The left image shows an example of a consecutive slide double stained with CD20 and CD3 IHC. The black square indicates the area that is shown in the central image with CD20^+^ B cells (in yellow) and CD3^+^ T cells (in purple) localizing in a TLS. The right image shows the digital markup with CD20-positive areas highlighted by the intensity of the yellow staining (depicted as spectrum from yellow to red color). **(E)** Example of a consecutive slide stained for PD-L1 IHC. The black square indicates the area that is shown in the right image. PD-L1 IHC slides were scored manually.

**Table 1.**
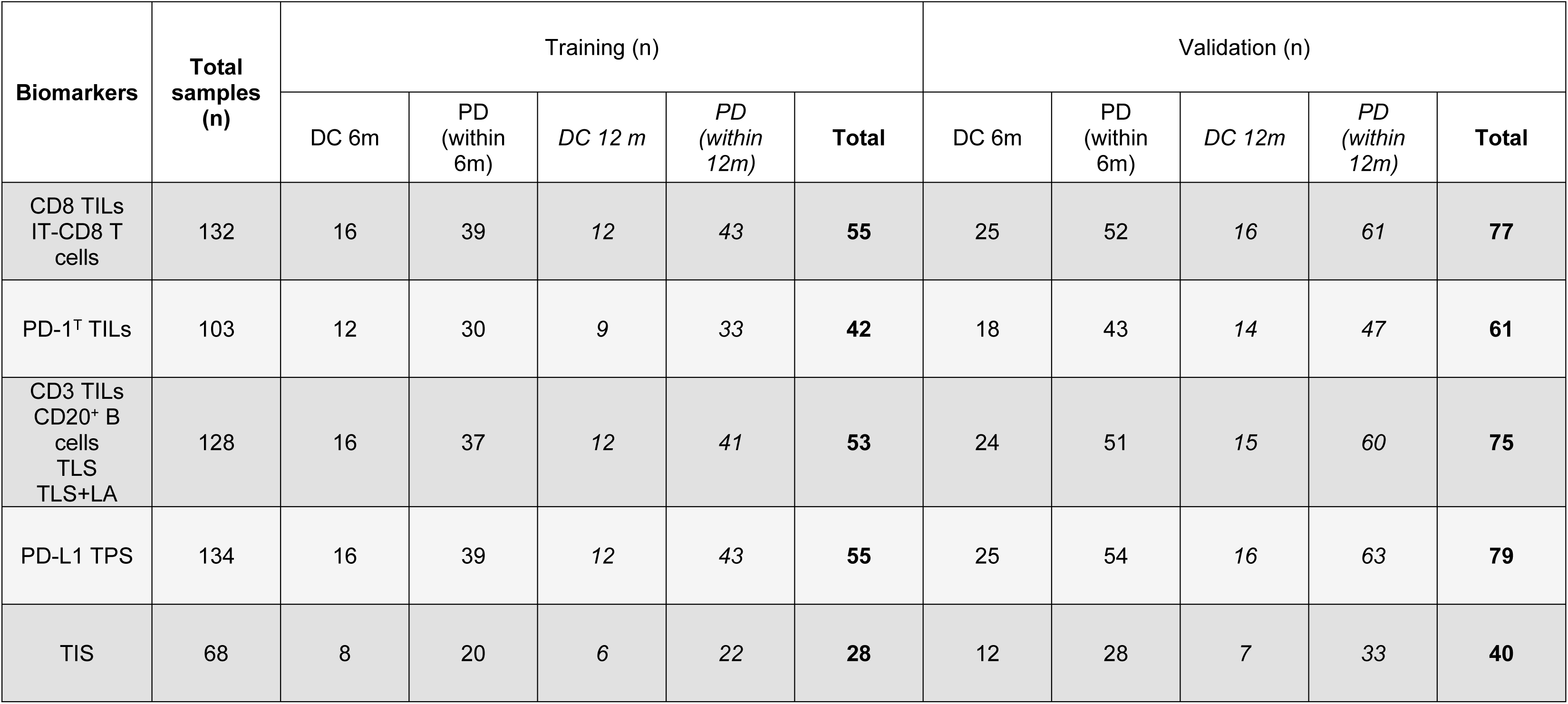
Total number of samples per biomarker in the training and validation cohort.

We randomized patients with ≥2 biomarker results available (n=135) in a training (n=55) and validation (n=80) cohort. This randomization was stratified for clinical benefit to ascertain that in both cohorts 1 in 3 patients reached DC 6m and 1 in 5 patients reached DC 12m, respectively. Since a limited number of patients with TIS scores (n=68) were available, these patients were randomly distributed proportionately **(Table 1, Supplementary Fig. S1)**. For every patient the results per biomarker are shown in **Supplementary Table S1**. Demographic characteristics did not significantly differ among the training and validation cohort **(Table 2)**.

**Table 2.**
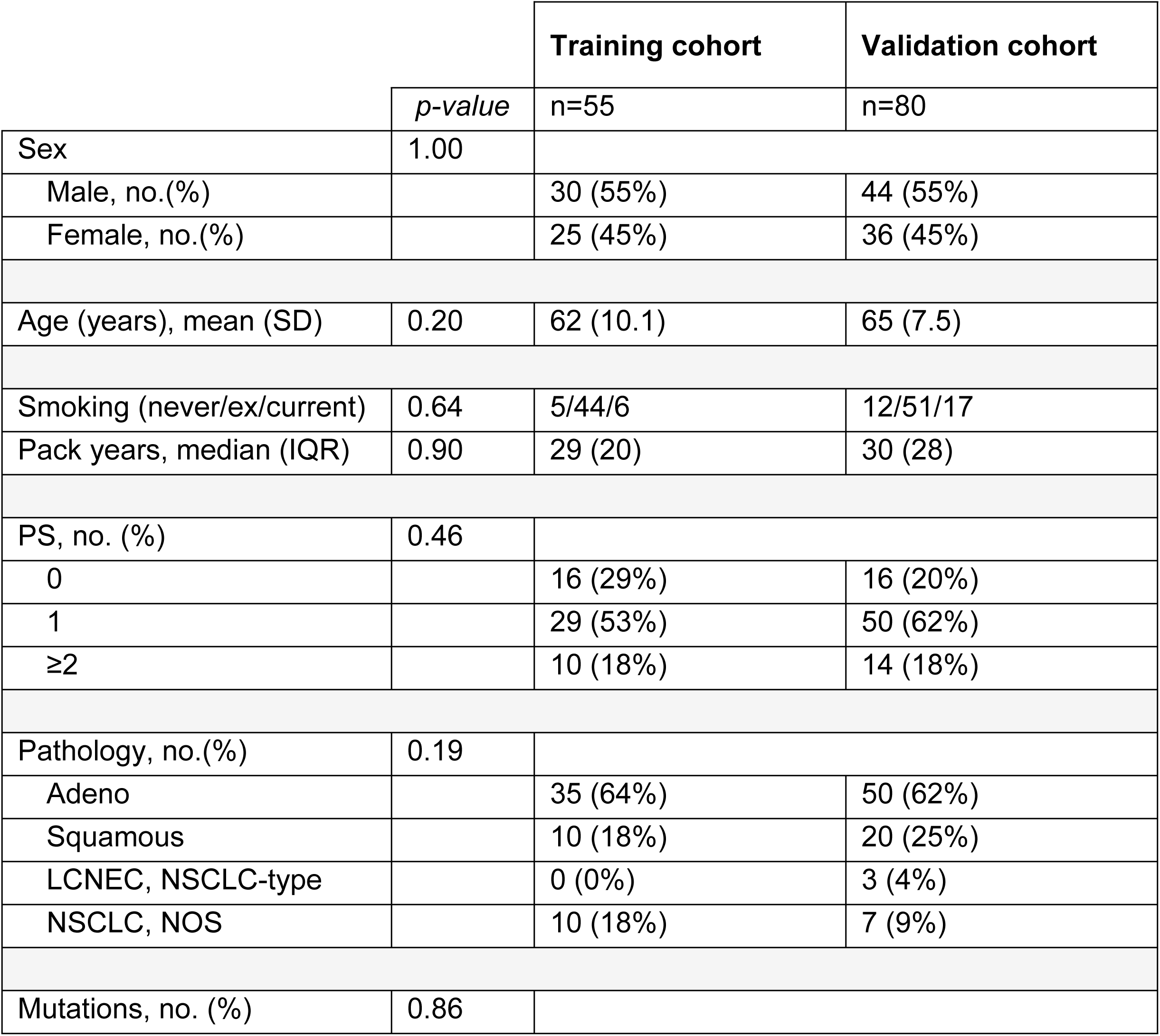
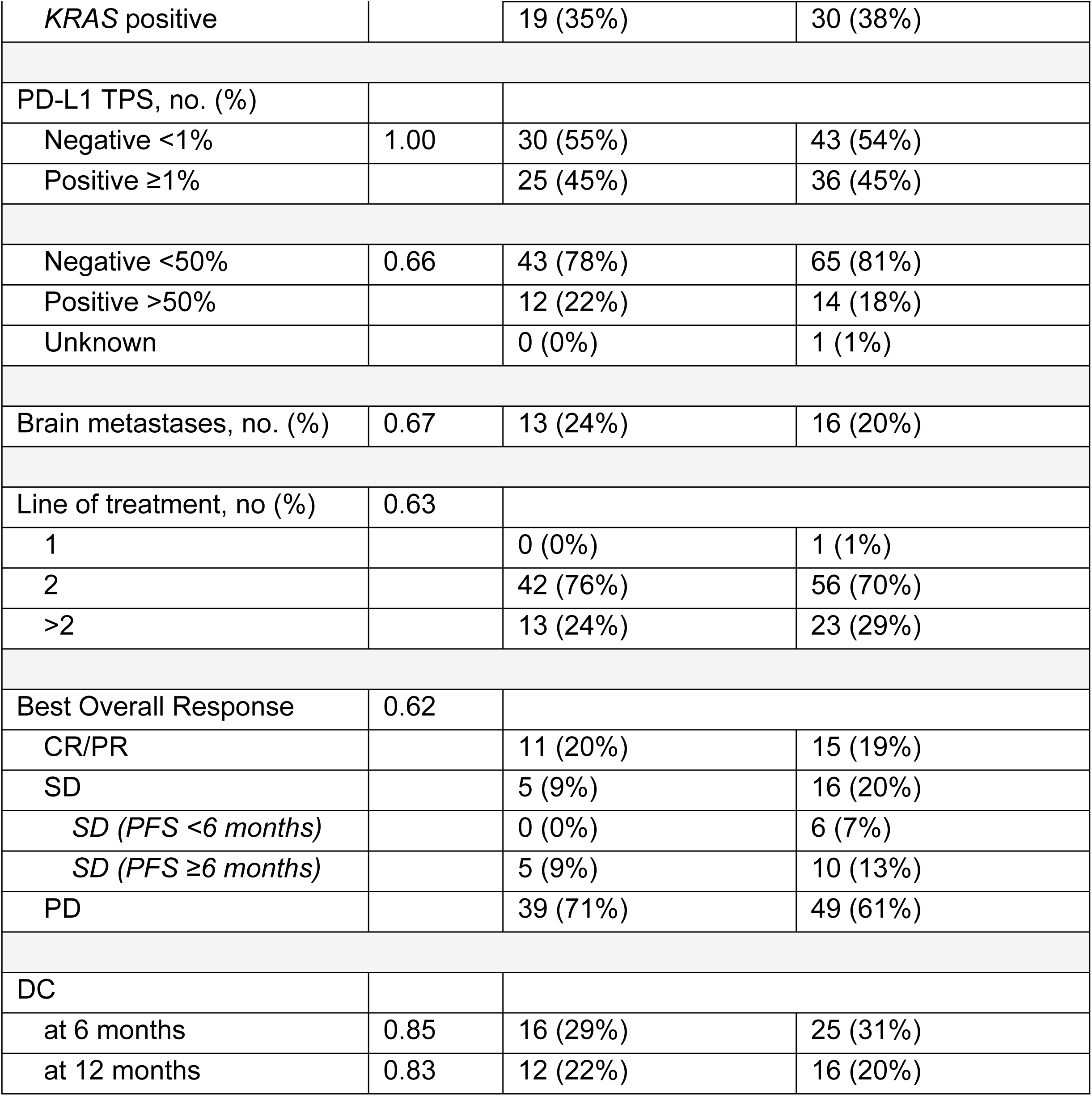
Patient characteristics and treatment outcomes for training and validation cohorts. *P*-values were calculated by Mann-Whitney, Fisher exact or linear-by-linear association tests. *S.d*, standard deviation; *IQR*, interquartile range; *PS*, Performance Score, based on the European Cooperative Oncology group (ECOG) performance status score. This is a score ranging from 0 to 5, where 0 indicates no symptoms, 1 indicates mild symptoms and above 1 indicates greater disability; *LCNEC NSCLC* type, large cell neuroendocrine carcinoma non-small cell lung cancer type; *NOS*, not otherwise specified; *KRAS*, Kirsten Rat Sarcoma viral oncogene; *PD-L1*, programmed death ligand 1; *TPS*, tumor proportion score; *CR*, complete response; *PR*, partial response; *SD*, stable disease; *PD*, progressive disease; *DC*, disease control.

### Accuracy of individual and composite biomarkers to predict DC at 6 months

Next, we determined the most optimal cut-offs for each individual and composite biomarkers in the training cohort. We aimed for a sensitivity and NPV of ≥90% to minimize undertreatment and a specificity of at least 50% to identify those patients that are unlikely to respond to PD-1 blockade therapy and can potentially benefit from alternative treatments. Since not all tumor samples were evaluable for all nine biomarkers, the number of training samples ranged from 28 to 55 **(Table 1, Supplementary Fig. S1)**. In total, 16 composite biomarkers and PD-1^T^ and TIS as individual biomarkers reached ≥90% sensitivity and ≥50% specificity **(Supplementary Table S2)**. Interestingly, these include 7/8 (88%) possible combinations with PD-1^T^ TILs and 5/8 (63%) with TIS **(Supplementary Table S2)**. However, none of these combinations did significantly improve predictive accuracy compared to PD-1^T^ TILs and TIS alone **(Supplementary Fig. S2A,B)** and were excluded from further analysis.

Next, we selected the four remaining biomarkers with the highest predictive performance for validation, being the combinations of CD8+IT-CD8 and CD3+IT-CD8, as well as PD-1^T^ TILs and TIS alone, respectively **(Supplementary Table S2)**. In the training cohort, both CD8+IT-CD8 and CD3+IT-CD8 had significantly higher probability scores in the DC 6m group (reflecting the probability of patients reaching DC 6m) compared to the PD group (CD8+IT-CD8, *P*<0.0001 and CD3+IT-CD8, *P*<0.001) **(Fig. 2A,B)**. The area under the ROC curve (AUC) was 0.83 (95% CI 0.73-0.94) for CD8+IT-CD8, and 0.78 (95% CI 0.65-0.92) for CD3+IT-CD8 **(Fig. 2C,D)**. Cut-offs of 0.167 and 0.161, respectively, correlated to a sensitivity of 94% and 94%, specificity of 62% and 54%, NPV of 96% and 95% and PPV of 50% and 47% **(Table 3)**.

**Figure 2.**
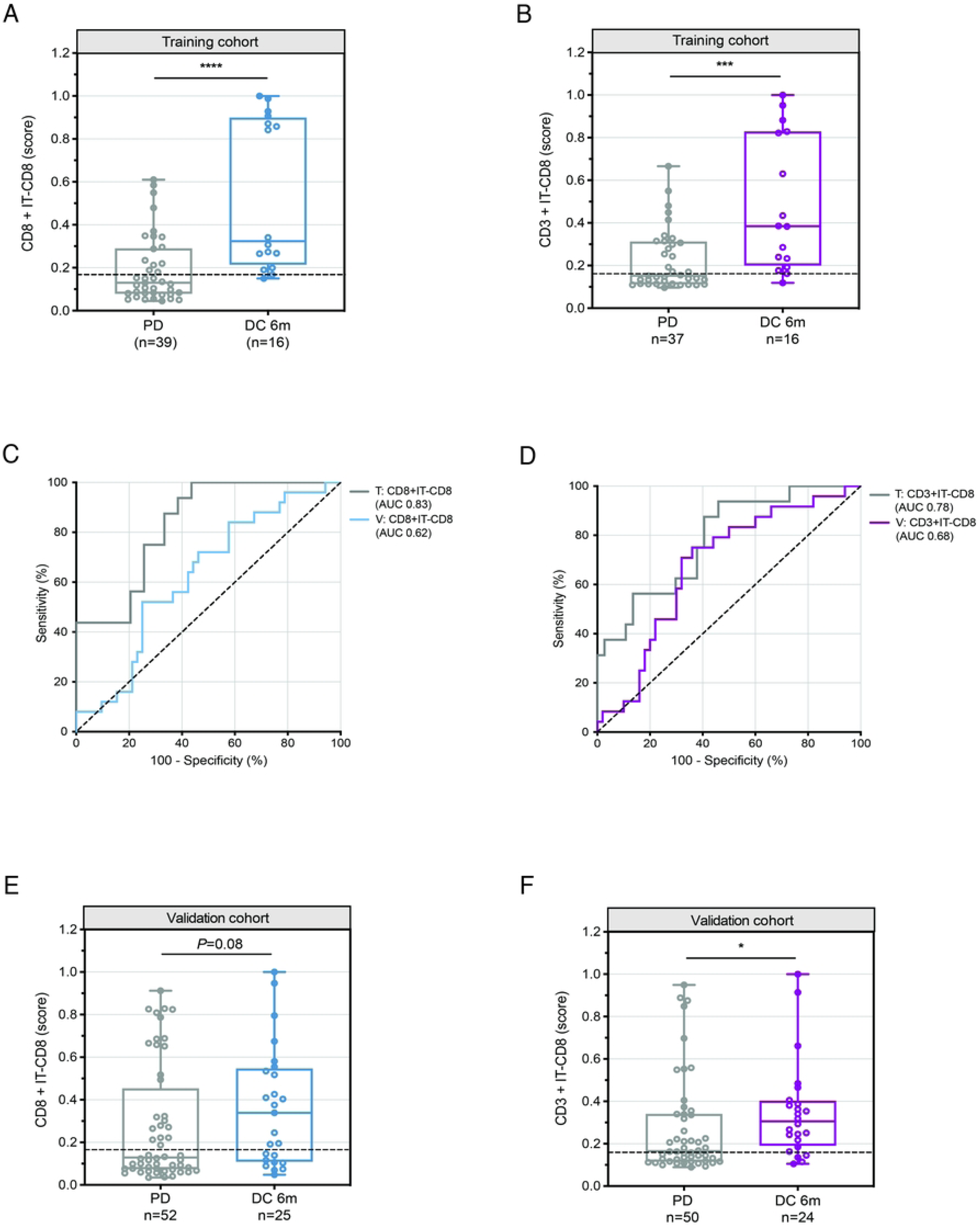
Performance of selected composite and individual biomarkers to predict DC at 6 months in NSCLC patients treated with PD-1 blockade. **(A)** Probability scores of CD8+IT-CD8 in pretreatment samples from patients with disease control at 6 months (DC 6m) (n=16) and progressive disease (PD) (n=39) in the training cohort (n=55). Dashed line indicates a cut-off of 0.167. Medians, interquartile ranges and minimum/maximum shown in boxplots, \*\*\*\**P*<0.0001 by Mann Whitney U-test. **(B)** Probability scores of CD3+IT-CD8 in pretreatment samples from patients with DC 6m (n=16) and PD (n=37) in the training cohort (n=53). Dashed line indicates a cut-off of 0.161. Medians, interquartile ranges and minimum/maximum shown in boxplots, \*\*\**P*<0.001 by Mann Whitney U-test. **(C)** Receiver operating characteristic (ROC) curve for predictive value of CD8+IT-CD8 for DC 6m in the training (n=55) (AUC 0.83, 95% CI: 0.73-0.94) and validation cohort (n=77) (AUC 0.62, 95% CI: 0.50-0.75). **(D)** ROC curve for predictive value of CD3+IT-CD8 for DC 6m in the training (n=53) (AUC 0.78, 95% CI: 0.65-0.91) and validation cohort (n=74) (AUC 0.68, 95% CI: 0.55-0.80). **(E)** Same plot as in **A** (CD8+IT-CD8) for patients with DC 6m (n=25) and PD (n=52) in the validation cohort (n=77), *P*=0.08. **(F)** Same plot as in **B** (CD3+IT-CD8) for patients with DC 6m (n=24) and PD (n=50) in the validation cohort (n=74), \**P*=0.02.

**Table 3.** Predictive accuracy of selected individual and composite biomarkers, summary of training and validation results.

|  |  |  |  |  | Training |  |  |  |  |  | Validation |  |  |  |  |  |
| --- | --- | --- | --- | --- | --- | --- | --- | --- | --- | --- | --- | --- | --- | --- | --- | --- |
| Clinical outcome | Bio-marker | Predictor | Cut-off | Samples (n) | AUC (95% CI) | Sensitivity | Specificity | NPV | PPV |  | Samples (n) | AUC (95%-CI) | Sensitivity | Specificity | NPV | PPV |
| DC 6 months | PD-1<sup>T</sup> TILs |  | 90 | 42 | 0.82 (0.69-0.95) | 92% | 67% | 95% | 52% |  | 61 | 0.72 (0.57-0.87) | 72% | 74% | 86% | 54% |
|  | TIS |  | 6.65 | 28 | 0.81 (0.65-0.98) | 100% | 55% | 100% | 47% |  | 40 | 0.57 (0.36-0.77) | 83% | 39% | 84% | 37% |
|  | CD8+ IT-CD-8 | probability for DC = 1 / (1 - exp (-3.5749 + 0.0031 * CD8 + 0.043 * IT-CD8)) | 0.167 | 55 | 0.83 (0.73-0.94) | 94% | 62% | 96% | 50% |  | 77 | 0.62 (0.50-0.75) | 64% | 56% | 76% | 41% |
|  | CD3+ IT-CD-8 | probability for DC = 1 / (1 - exp (-2.3821 + 0.0806 * CD3 + 0.0175 * IT-CD8 + 0.0069 * CD3 * IT-CD8)) | 0.161 | 53 | 0.78 (0.65-0.91) | 94% | 54% | 95% | 47% |  | 74 | 0.68 (0.55-0.80) | 83% | 46% | 85% | 43% |
| DC 12 months | PD-1<sup>T</sup> TILs |  | 90 | 42 | 0.82 (0.70-0.94) | 100% | 64% | 100% | 43% |  | 61 | 0.80 (0.65-0.94) | 86% | 74% | 95% | 50% |
|  | TIS |  | 6.65 | 28 | 0.77 (0.58-0.96) | 100% | 50% | 100% | 35% |  | 40 | 0.63 (0.43-0.82) | 100% | 39% | 100% | 26% |
|  | CD8+ IT-CD-8 | probability for DC = 1 / (1 - exp (-4.0644 + 0.003 * CD8 + 0.0436 * IT-CD8)) | 0.122 | 55 | 0.85 (0.73-0.96) | 92% | 63% | 96% | 41% |  | 77 | 0.67 (0.53-0.81) | 68% | 57% | 88% | 30% |
|  | CD8+ TIS | probability for DC = 1 / (1 - exp (-5.7952 + 0.0224 * CD8 + 0.2346 * TIS + -0.0021 * CD8 * TIS)) | 0.124 | 28 | 0.91 (0.79-1.00) | 100% | 68% | 100% | 46% |  | 38 | 0.59 (0.36-0.82) | 29% | 68% | 81% | 17% |

Also, the PD-1^T^ TIL numbers and TIS scores were significantly higher in the DC 6m group than in the PD group (PD-1^T^ TILs, *P*<0.001 and TIS, *P*<0.01) **(Supplementary Fig. S2C,D)**. PD-1^T^ TILs showed an AUC of 0.82 (95% CI 0.69-0.95) and TIS an AUC of 0.81 (95% CI 0.65-0.98) **(Supplementary Fig. S2E,F)**. For PD-1^T^ TILs a cut-off of 90 per mm^2^ was chosen, as this cut-off showed predictive value in a prior study(25). We observed a sensitivity of 92%, specificity of 67%, NPV of 95% and PPV of 52% **(Table 3)**. A score of 6.65 was chosen as optimal cut-off for TIS which correlated to a sensitivity of 100%, specificity of 55%, NPV of 100% and PPV of 47% **(Table 3)**.

Next, we evaluated the predictive performance of the four selected biomarkers in the validation cohort. The number of validation samples with successful biomarker results ranged from 40 to 79 **(Table 1, Supplementary Fig. S1)**. We observed that the predictive accuracy of CD8+IT-CD8 and CD3+IT-CD8 biomarkers was substantially lower compared to the training cohort. Specifically, probability scores in the DC 6m group did not significantly differ from scores in the PD group for CD8+IT-CD8 (*P*=0.08) **(Fig. 2E).** For CD3+IT-CD8 this comparison was borderline significant (*P*=0.01) **(Fig. 2F)**. The AUC of the ROC curve was 0.62 (95% CI 0.50-0.75) for CD8+IT-CD8 and 0.68 (95% CI 0.55-0.80) for CD3+IT-CD8 **(Fig. 2C,D)**. CD8+IT-CD8 reached a sensitivity of 64%, specificity of 56%, NPV of 76% and PPV of 41%. The predictive accuracy of CD3+IT-CD8 was higher than CD8+IT-CD8 but still lower than in the training cohort, reaching a sensitivity of 83%, specificity of 46%, NPV of 85% and PPV of 43% **(Table 3)**.

The individual biomarkers in the validation cohort showed that PD-1^T^ TIL numbers were significantly higher in the DC 6m group versus the PD group (*P*<0.01), which was not observed for TIS scores (*P*=0.52) **(Supplementary Fig. S2G,H)**. The discriminatory ability of PD-1^T^ TILs was lower as in the training, but still reached an AUC of 0.72 (95% CI 0.57-0.87) **(Supplementary Fig. S2E)**. TIS reached an AUC of 0.57 (95% CI 0.36-0.77) **(Supplementary Fig. S2F)**. A cut-off of 90 PD-1^T^ TILs per mm^2^ correlated to a sensitivity of 72%, specificity of 74%, NPV of 86% and PPV of 54%. A TIS score of 6.65 showed a comparable sensitivity (83%), NPV (84%) and PPV (37%) but lower specificity (39%) **(Table 3)**. In summary, these results demonstrate that a combination of CD8+IT-CD8 and CD3+IT-CD8 did not improve predictive accuracy compared to PD-1^T^ TILs and TIS alone. Furthermore, none of the selected biomarkers reached the prespecified performance criteria.

### Accuracy of individual and composite biomarkers to predict DC at 12 months

Approximately 70-80% of patients treated in 2^nd^ line with PD-(L)1 blockade progress within 12 months(2–4). We previously demonstrated that PD-1^T^ TILs could more effectively identify patients with DC 12m as compared to DC 6m, as well as a patient group without long-term benefit(25). We therefore also assessed the performance of all biomarkers to predict DC 12m. Similar to the DC 6m analysis, we determined the most optimal cut-offs for each of the composite and individual biomarkers to identify patients with DC 12m and with PD. Four patients in the training and nine patients in the validation experienced disease progression between 6 and 12 months, and were therefore included in the PD group in this analysis. 16 composite biomarkers reached ≥90% sensitivity and ≥50% specificity in the training cohort, as well as PD-1^T^ TILs and TIS as individual biomarkers **(Supplementary Table S3)**. We observed that 12/16 composite and 2/2 individual biomarkers (PD-1^T^ TILs and TIS), matched the 6-months endpoint with similar accuracy **(Supplementary Table S2,3)**. PD-1^T^ TIL combinations did not significantly improve predictive accuracy compared to PD-1^T^ TILs alone and were excluded from further analysis **(Supplementary Fig. S3A, Supplementary Table S3)**. However, the combination of CD8 with TIS (CD8+TIS) showed an increase of 18% specificity compared to TIS alone. This combination was selected for further analysis, even though it did not reach statistical significance, possibly due to the low sample size (*P*=0.34) **(Supplementary Fig. S3B, Supplementary Table S3)**. The four biomarkers with the highest predictive performance were selected for validation. 3/4 selected biomarkers, including PD-1^T^ TILs, TIS and CD8+IT-CD8, matched the DC 6m selection. The fourth biomarker included the combination of CD8+TIS **(Supplementary Table S3)**.

The probability scores for DC 12m and PD are shown per sample in **Supplementary Fig. S3C** (CD8+IT-CD8, *P*<0.001) and **Supplementary Fig. S3D** (CD8+TIS, *P*<0.01). The two composite biomarkers showed a high AUC of 0.85 (95% CI: 0.73-0.96) (CD8+IT-CD8) and 0.91 (95% CI: 0.79-1.00) (CD8+TIS) in the training cohort **(Fig. 3A,B)**. A cut-off of 0.122 and 0.124, respectively, was chosen as optimal cut-off **(Table 3)**. PD-1^T^ TIL numbers and TIS scores are shown in **Supplementary Fig S3E** and **F**. A cut-off of 90 PD-1^T^ TILs per mm^2^ and a TIS score of 6.65 demonstrated similar predictive accuracy as in the training for DC 6m **(Fig. 3C,D**, **Table 3)**.

**Figure 3.**
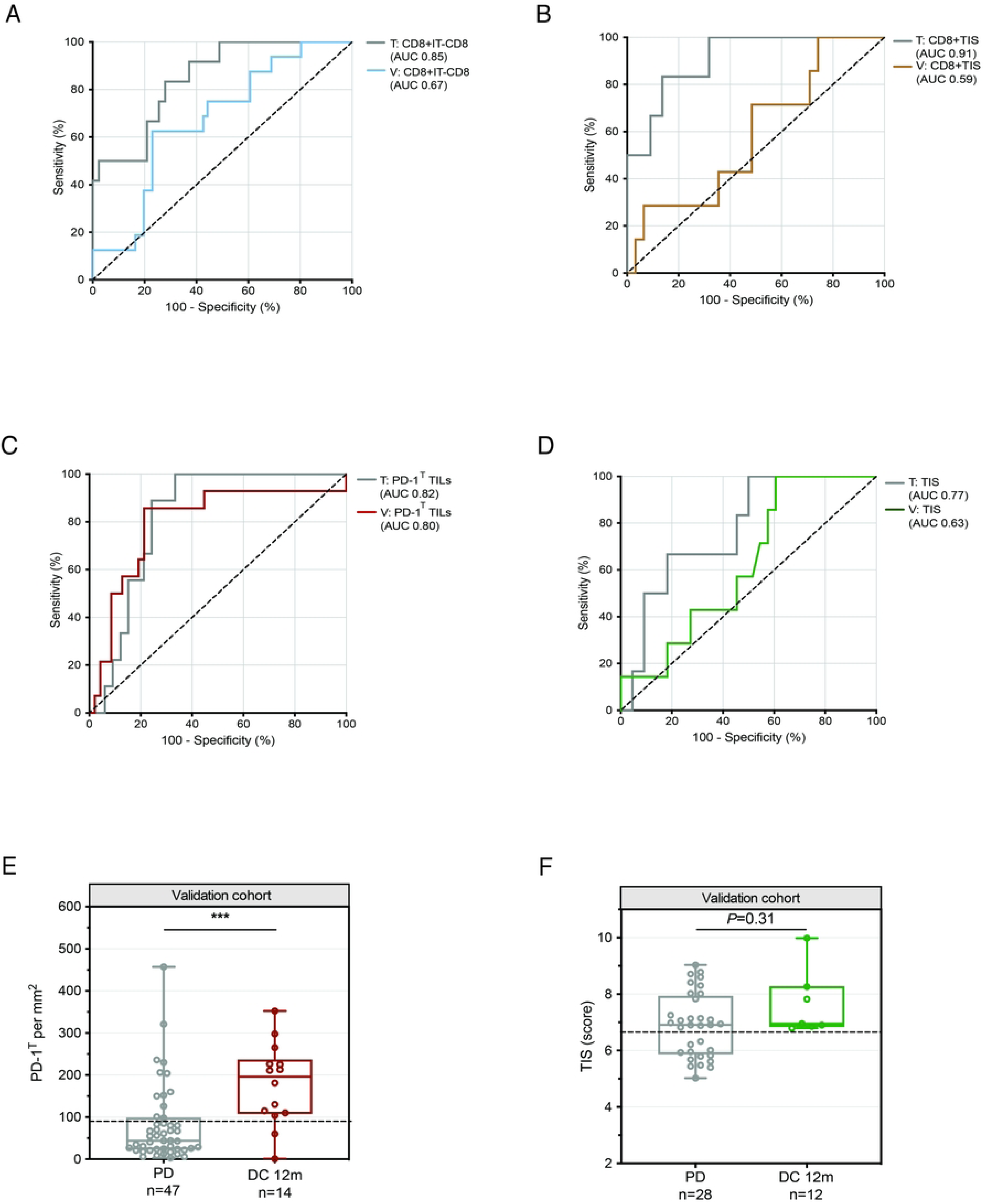
Performance of selected composite and individual biomarkers to predict DC at 12 months in NSCLC patients treated with PD-1 blockade. **(A)** Receiver operating characteristic (ROC) curve for predictive value of CD8+IT-CD8 for disease control at 12 months (DC 12m) in the training cohort (n=55) (AUC 0.85, 95% CI: 0.73-0.96) and validation cohort (n=77) (AUC 0.67, 95% CI: 0.53-0.81). **(B)** ROC curve for predictive value of CD8+TIS for DC 12m in the training cohort (n=28) (AUC 0.91, 95% CI: 0.79-1.00) and validation cohort (n=38) (AUC 0.59, 95% CI: 0.36-0.82). **(C)** ROC curve for predictive value of PD-1^T^ TILs for DC 12m in the training cohort (n=42) (AUC 0.82, 95% CI: 0.70-0.94) and validation cohort (n=61) (AUC 0.80, 95% CI: 0.65-0.94). **(D)** ROC curve for predictive value of TIS for DC 12m in the training cohort (n=28) (AUC 0.77, 95% CI: 0.58-0.96) and validation cohort (n=40) (AUC 0.63, 95% CI: 0.43-0.82). **(E)** PD-1^T^ TILs per mm^2^ in pretreatment samples from patients with DC 12m (n=14) and PD (n=47) in the validation cohort (n=61). Dashed line indicates a cut-off of 90 PD1^T^ TILs per mm^2^. Medians, interquartile ranges and minimum/maximum shown in boxplots, \*\**P*<0.01 by Mann Whitney U-test. **(F)** TIS scores in pretreatment samples from patients with DC 12m (n=7) and PD (n=33) in the validation cohort (n=40). Dashed line indicates a cut-off score of 6.65. Medians, interquartile ranges and minimum/maximum shown in boxplots, *P*=0.31 by Mann Whitney U-test.

In the validation cohort, of the two composite biomarkers, only CD8+IT-CD8 showed borderline significantly higher probability scores in the DC 12m group versus the PD group (*P*=0.03) **(Supplementary Fig. S3G,H)**. The ROCs yielded low AUCs (CD8+IT-CD8: 0.67 (95% CI: 0.53-0.81), CD8+TIS: 0.59 (95% CI 0.36-0.81)) **(Fig 3A,B)**. Furthermore, the sensitivity (68% and 29%), specificity (57% and 68%), NPV (88% and 81%) and PPV (30% and 17%) did not meet the prespecified performance criteria **(Table 3)**.

PD-1^T^ TIL numbers were significantly higher in patients with DC 12m versus PD (*P*<0.001) **(Fig. 3E)**. PD-1^T^ TILs also demonstrated a consistently high AUC (0.80, 95% CI: 0.65-0.94) and high accuracy, reaching a sensitivity of 86%, specificity of 74%, NPV of 95% and PPV of 50% **(Fig. 3C**, **Table 3)**. We observed an enrichment of patients with DC 12m in the ≥90 group and with PD in the <90 subgroup **(Fig. 3E)**. TIS scores did not significantly differ between the two groups (*P*=0.31) and showed a low AUC of 0.63 (95% CI 0.43-0.82) **(Fig. 3D,F)**. However, a cut-off score of 6.65 reached a sensitivity of 100%, specificity of 39%, NPV of 100% and PPV of 26% **(Table 3)**. These findings did not meet our ≥50% specificity criterium, but accurately identified all patients with DC 12m including 39% of patients with PD **(Fig. 3F)**. Taken together, PD-1^T^ TILs and TIS as individual biomarkers showed higher predictive accuracy for DC 12m compared to the combination of CD8+IT-CD8 and CD8+TIS. Notably, PD-1^T^ TILs alone was more accurate than TIS alone, as specificity and PPV were substantially higher.

## Discussion

Since the introduction of PD-(L)1 blockade therapy, clinical outcome of advanced stage NSCLC has dramatically improved. Nevertheless, a subset of patients derive benefit from these treatments which consequently has led to overtreatment and unnecessarily side effects in many. In addition, health care systems deal with increasing costs. Several predictive biomarkers have been identified to support treatment decision making. Since different components in the TME can affect the tumor immune response upon PD(L)1 blockade therapy, it is unlikely to find one single perfect biomarker. Based on the hypothesis that a predictive model should contain more than one biomarker; we here assess the predictive performance of biomarker combinations in an advanced stage NSCLC cohort treated with nivolumab. Our data showed that selected composite biomarkers did not improve predictive performance as compared to PD-1^T^ TILs and TIS alone. At 6 months, none of the selected composite and individual biomarkers reached the prespecified performance criteria in the validation cohort. At 12 months, PD-1^T^ TILs and TIS could identify patients with DC 12m with high accuracy. Patients without long-term benefit were more accurately identified by PD-1^T^ TILs than TIS.

Whereas CD8 or CD3 TILs in combination with intratumoral localization of CD8 T cells were the most accurate composite biomarkers for DC 6m in the training cohort, we observed that discriminatory ability was low in the validation cohort. The presence and localization of TILs alone might not indicate that all T cells are in a state to recognize and eliminate the tumor(36,37). In the present study, this notion is supported by the high accuracy of PD-1^T^ TILs to predict DC 12m, as these TILs have been identified as a distinct TIL subset with a high capacity of tumor recognition(24). The results are similar to our previous work because the majority of samples were re-used(25). Further refinement of this T cell population could contribute to the development of new markers or gene signatures, as recently been done by other studies(38–40). Since most of the biomarkers assessed in this study are related to antitumor immunity and are presumably correlated, PD-1^T^ combinations did not improve specificity.

Previous studies have shown the predictive potential of combining CD8+PD-L1(22,34). However, in our training cohort, CD8+PD-L1 did not meet our performance criteria, and as a result, this combination was not further evaluated. Noguchi et al. previously observed that PD-L1 expression on tumor cells is transient and dependent on the production of IFNγ by TILs(41). Hence, variable tumor PD-L1 expression in training samples might have affected the predictive accuracy of PD-L1 alone and that of PD-L1 combinations. Furthermore, this study is limited by the number of samples, in particularly for TIS assessment. Therefore, we restricted our evaluation to two-biomarker combinations instead of considering three or more. Studies involving a larger number of samples are essential to further validate our findings.

Our results for TIS are in line with other studies that demonstrated the predictive potential of this signature(29,42). Interestingly, TIS contains genes that are highly expressed in PD-1^T^ TILs, such as LAG3 and TIGIT(24,29). A high number of PD-1^T^ TILs or a high TIS score in pretreatment samples may serve as surrogate markers for a tumor’s ability to undergo durable immune reactivation upon PD-1 blockade therapy. A PD-1^T^ TILs or TIS combination with biomarkers reflecting distinct parts of the immune response could potentially improve predictive accuracy. For example, TMB can serve as a read-out for immunogenic neoantigens that arise from somatic mutations(15). TMB and PD-L1 have previously been described as independent predictors for advanced NSCLC treated with PD-1 blockade and have shown improved performance when combined(32,33). Another suggestion is, in contrast, the presence of tumor-resident regulatory T cells (T_reg_) in the TME. T_reg_ cells possess an immune-inhibitory function and high numbers are correlated to poor patient survival(43). A combination of TMB or T_reg_ with either PD-1^T^ or TIS could be further explored in future work.

In conclusion, this study showed that the biomarker combinations assessed here did not improve predictive performance when compared to PD-1^T^ TILs and TIS alone. PD-1^T^ TILs showed the highest predictive performance of all biomarkers, as patients with no long-term benefit were identified with high specificity and NPV.

## Data Availability

All relevant data are within the manuscript and its Supporting Information files.

## Acknowledgements

We would like to thank the NKI-AVL Core Facility Molecular Pathology and Biobanking for supplying all IHC stainings used in this study, as well as biobank-related work and other laboratory support.

